# Disparities in research capacity for tobacco control: an inventory of peer-reviewed publications using the Global Tobacco Surveillance System data

**DOI:** 10.1101/2022.09.08.22279271

**Authors:** Isabel Garcia de Quevedo, Amulya Marellapudi, Edward Rainey, Evelyn Twentyman

## Abstract

Peer-reviewed publications using tobacco surveillance data represent a critical step toward evidence-based tobacco control, but research and publication capacity in countries with fewer resources may be limited. This paper describes patterns in use of the Global Adult Tobacco Survey (GATS) and/or Global Youth Tobacco Survey (GYTS) data for publications, investigates the origin of the data versus the origin of lead authorship, and describes geographic patterns of publications by country, region, and country income. A bibliometric inventory using six search engines was conducted for relevant studies using data from either of these surveys between January 1999 and January 2021. Descriptive statistics, including frequencies and percentages,were used to analyze publication characteristics. Our search strategy retrieved 1,834 initial records; 493 publications were ultimately included: 249 (50.5%) used adult surveillance data (GATS) and 248 (50.3%) used youth surveillance data (GYTS). Most publications were in English (97.2%, n=479). Data published 50 or more times represented 12 low- and middle-income countries (LMIC): India, Bangladesh, Vietnam, Mexico, Egypt, Thailand, Poland, Philippines, China, Russian Federation, Turkey and Ukraine. While many of the papers analyzed data from low- and middle-income countries, the number of publications by origin of lead author was the highest for the United States (*n*=135) and India (*n*=84). Over 80% of the world’s 1.3 billion tobacco users live in low- and middle-income countries (LMIC) and we found underrepresentation of these countries as lead authors. These findings can be used to identify opportunities to enhance capacity for analysis, research and dissemination of global tobacco control data in LMIC.

## Introduction

Enhancing research capacity, defined as the ability to engage in, perform or carry out quality research, has the potential to support policies and programs and hence prioritize public health issues (1, 2). Strengthening research capacity and dissemination for global tobacco control and scientific publications is critical to implement multisector policies and programs, especially in low- and middle-income countries (LMIC) where the majority of the burden lies(3-6). The incongruence between the high burden of diseases in LMIC, and low research productivity has been well documented (7-9). Authors from LMIC have disproportionately fewer publications than authors in HIC (10, 11) while, conversely, most of the burden of tobacco use falls on LMIC, which often have fewer financial resources and understaffed healthcare and academic workforce(1, 2). The decline in tobacco use for high income countries has been attributed in part to coordinated efforts guided by a strong research agenda, population-based surveillance, and communication of local research in scientific publications, all of which has driven implementation of programs and policies (3).

Tobacco kills more than eight million people each year worldwide, including 1.2 million people from secondhand smoke (12). Over 80% of people who smoke reside in LMIC, which are disproportionately affected by the burden of tobacco use and this in turn places a large burden on governments, health systems and individuals (13-15). In addition, there is a stark mismatch between the location where these peer-reviewed papers come from and the burden that affects LMIC, especially studies that focus on non-communicable diseases and its risk factors, including tobacco use (7, 16-18).

The World Health Organization Framework Convention on Tobacco Control (WHO FCTC) (19), was the first international treaty that provided key measures to implement and manage tobacco control. The FCTC has been critical since its inception in 2003, for signatory countries to have a research agenda to effectively monitor tobacco control regionally and locally. The Global Tobacco Surveillance System (GTSS) was established in 1998 to enhance the capacity of countries to design, implement and evaluate their national comprehensive tobacco action plan, and, for signatories, to monitor the key articles of the WHO FCTC (20). Two of GTSS’ main components: The Global Youth Tobacco Survey (GYTS) started implementation on 1999 and the Global Adult Tobacco Survey (GATS) started in 2008. To date, GTSS has assisted over 180 countries and locations to implement at least one survey of tobacco use and key tobacco control measures; many countries have completed multiple surveys and multiple rounds of the same survey (21, 22).

Twenty years after GTSS started, we still do not have an objective measurement of the scientific output for tobacco surveillance data. It is not yet clear how or whether the data collected through these surveys have been incorporated into research agendas leading to implementation of relevant programs and policies. There is no compendium or inventory of where these data were published, by whom these data have been published, and whether these data are published by lead authors coming from the country in which these data were collected. Such an inventory can be a step in the first determination of how or whether the data collected through these surveys have been incorporated into research agendas leading to implementation of relevant programs and policies. In this review, we present a bibliometric inventory of peer reviewed scientific publications that use two specific GTSS components: The GYTS and GATS. This inventory will help determine whether, how and where published tobacco surveillance data has been used and will aim to identify research gaps and opportunities.

The objectives of this study were to 1) have a bibliometric inventory of peer-reviewed publications that used GTSS data, 2) to investigate the origin of the data versus the origin of lead authorship, 3) investigate geographic patterns of publications by country, region, and income, and 4) investigate patterns in use of GTSS data for publications.

## Methods

### Search Strategy

We searched six databases (Embase, MEDLINE including Epub ahead of print, Scopus, Scielo, BDSP and World Health Organization Regional Databases - Global Index Medicus) for relevant studies that included GTSS data between 1999 and January 2021. We restricted the search to publications in English, Spanish, French or Chinese. We included the following search terms: “tobacco AND Global Adult Tobacco Survey OR Global Youth Tobacco Survey OR GATS OR GYTS”

### Eligibility Criteria

We included publications that: (1) used GATS and/or GYTS data in their methodology, (2) were a research article or abstract with the outcome being tobacco use (3) were in English, Spanish, French or Chinese.

We excluded 21 publications that mentioned in their methods they used an adapted or unofficial version of GATS or GYTS questionnaire in their methodology.

### Data Selection and Extraction

We screened the records based on titles and abstracts retrieved from the search. For publications that were identified as potentially relevant, we retrieved and reviewed the full text. We also retrieved the full text if the abstract and title were not enough to decide eligibility. Two researchers extracted data from the potential publications. We used a predefined data extraction matrix to collect data from all eligible studies. The data extraction matrix was adapted from the Preferred Reporting Items for Systematic Reviews and Meta-Analysis (PRISMA) 2020 checklist (23). The information from non-English papers was extracted and entered into the database in English by two researchers that spoke the language. There were no French papers that met the eligibility criteria for extraction.

Data extracted included: year of publication, type of publication (manuscript or abstract), language of publication (English, Spanish, Chinese or French), dataset used (GATS or GYTS), origin of GTSS data by country (ies) and by WHO region and World Bank country income classification, years of GATS/GYTS conducted, lead author country of origin (defined by lead author’s affiliation institution on the publication), international collaboration (defined by co-authors’ affiliations), focus of the publication on one or more of the WHO MPOWER measures (24), type of tobacco analyzed in the article (smoked tobacco, smokeless tobacco, or e-cigarettes), age of study participants, sex of study participants, summary of methods, study outcome variable(s), main finding/summary of study’s results, statistical program used for the analysis and use of other dataset(s) in addition to GATS or GYTS.

### Data Analysis

To look at the characteristics for each GTSS publication, we tabulated the publications in each category using descriptive statistics, including frequencies and percentages. Publication trends were analyzed by year and type of data set used (GATS or GYTS).

After extracting data for lead author’s origin and origin of GTSS data, we grouped countries based on the WHO regions: WHO Regional office for Africa (WHO/AFRO) (25),WHO Regional office for Europe (WHO/EURO) (26), WHO Regional Office for the Eastern Mediterranean (WHO/EMRO) (27),Regional office for the South-East Asia Region (WHO/SEARO) (28),Pan American Health Organization (WHO/PAHO) (29) and WHO regional Office for the Western Pacific Region (WHO/WPRO) (30), and by country income level according to the 2021 World Bank Income classification criteria which classifies countries as: high income countries (h-ICs), upper middle income countries (u-MICs), lower middle income countries (l-MICs); and low income countries (l-ICs) (31). For context, we qualitatively observed the relationship between the number of country specific research papers and the country’s adult tobacco use burden by juxtaposing both indicators (adult tobacco use versus number of research papers per country).

We screened lead authorship data for publication outliers, which were defined as values greater than three standard deviations from the mean. Authorship in two countries – India and US – were outliers and were therefore analyzed independently from their respective region.

## Results

Our search strategy resulted in 1,834 records (532 from Medline, 655 from Embase, 586 from Scopus, 51 from Scielo and 10 from BDSP). After removing duplicates (n=976), we screened 858 records and further excluded 306 due to the abstract and/or title not meeting the inclusion criteria (1. used GATS and/or GYTS data in their methodology, 2. were a research article or abstract with the outcome being tobacco use, 3. were in English, Spanish, French or Chinese). That left us with a total of 555 publications for final extraction. After reviewing the full text publication, we deemed 62 articles ineligible because they didn’t meet the inclusion criteria. A final count of 493 publications met the inclusion criteria and were included in our review (Fig 1).

### Characteristics of GTSS Publications

From the 493 publications reviewed, 251 (50.9%) used adult data (GATS) and 248 (50.3%) used youth data (GYTS) (Table 1). Six out of the 493 publications combined GATS and GYTS within the same publication. Fifty-two publications (10.5%) were abstracts for conferences and the rest were full manuscripts (80.5%, n=441). Most of the publications were in English (97.2%, n=479) and only a small number were in Spanish (2.2%, n=11) or Chinese (0.6%, n=3). We did not find any eligible publications in French. We looked at number of publications overtime since each of the surveys was implemented (GYTS started implementation in 1998 and GATS in 2008) and found no differences overtime.

**Table 1:** Characteristics of GTSS-related publications by survey, 1999-2021.

| Characteristic | GTSS<sup>a</sup> survey |  |  |
| --- | --- | --- | --- |
|  | Overall<br>n (%) | GATS<sup>b</sup><br>n (%) | GYTS<sup>c</sup><br>n (%) |
| Total number of publications<sup>d</sup> | 493 (100) | 251 (50.9) | 248 (50.3) |
| **Publication type<sup>d</sup>** |  |  |  |
| Manuscript | 441 (89.5) | 215 (85.7) | 232 (93.5) |
| Abstract only<sup>e</sup> | 52 (10.5) | 36 (14.3) | 16 (6.5) |
| **Publication language<sup>f</sup>** |  |  |  |
| English | 479 (97.2) | 245 (97.6) | 240 (96.8) |
| Chinese | 3 (0.6) | 2 (0.8) | 1 (0.4) |
| Spanish | 11 (2.2) | 4 (1.6) | 7 (2.8) |
| **Origin of GTSS data by WHO Region<sup>g</sup>** |  |  |  |
| AFRO | 97 (12.0) | 29 (6.9) | 69 (17.2) |
| EMRO | 106 (13.1) | 46 (11.0) | 62 (15.4) |
| EURO | 134 (16.6) | 66 (15.7) | 71 (17.7) |
| PAHO | 120 (14.8) | 70 (16.7) | 52 (12.9) |
| SEARO | 210 (26.0) | 136 (32.4) | 77 (19.2) |
| WPRO | 142 (17.6) | 73 (17.4) | 71 (17.7) |
| **Lead author by WHO region** |  |  |  |
| AFRO | 35 (7.1) | 9 (3.6) | 26 (10.5) |
| EMRO | 25 (5.1) | 7 (2.8) | 18 (7.3) |
| EURO | 76 (15.4) | 32 (12.9) | 45 (18.1) |
| PAHO - total | 183 (37.1) | 88 (35.3) | 96 (38.7) |
| PAHO - (without US) | 48 (9.7) | 24 (9.6) | 24 (9.7) |
| US alone | 135 (27.4) | 64 (25.7) | 72 (29.0) |
| SEARO – total | 107 (21.7) | 81 (32.5) | 27 (10.9) |
| SEARO (without India) | 22 (4.5) | 10 (4.0) | 12 (4.8) |
| India alone | 85 (17.2) | 72 (28.9) | 15 (6.0) |
| WPRO | 67 (13.6) | 31 (12.4) | 36 (14.5) |
| **Lead author by world bank country income classification<sup>h</sup>** |  |  |  |
| Low income | 9 (1.8) | 1 (0.4) | 8 (3.2) |
| Lower-middle income | 156 (31.6) | 106 (42.6) | 53 (21.4) |
| Upper-middle income | 99 (20.2) | 45 (18.1) | 54 (21.9) |
| High income | 229 (46.5) | 97 (39.0) | 133 (53.6) |
| **Type of tobacco** |  |  |  |
| Smoked tobacco use<sup>i</sup> | 466 (94.5) | 230 (92.4) | 240 (96.8) |
| Smokeless tobacco use<sup>j</sup> | 151 (30.6) | 99 (39.8) | 55 (22.2) |
| E-cigarette | 20 (4.1) | 9 (3.6) | 12 (4.8) |
| **Publications by sex** |  |  |  |
| Male only | 4 (0.8) | 3 (1.2) | 1 (0.4) |
| Female only | 10 (2.0) | 9 (3.6) | 2 (0.8) |
| Both (male & female combined) | 479 (97.2) | 237 (95.2) | 245 (98.8) |
| **Statistical software used for analysis** |  |  |  |
| Epi Info | 8 (1.6) | 3 (1.2) | 6 (2.4) |
| SAS | 35 (7.1) | 14 (5.6) | 21 (8.5) |
| SPSS | 108 (21.9) | 64 (25.7) | 44 (17.7) |
| STATA | 99 (20.1) | 51 (20.5) | 48 (19.4) |
| SUDAAN | 53 (10.8) | 3 (1.2) | 51 (20.6) |
| STATISTICA | 15 (3.0) | 13 (5.2) | 2 (0.8) |
| R | 6 (1.2) | 2 (0.8) | 4 (1.6) |
| Other<sup>k</sup> | 21 (4.6) | 11 (4.4) | 10 (4.0) |
| Not reported | 148 (30) | 88 (35.3) | 62 (25) |

NOTE: “Origin of GTSS data by WHO region” and “Type of Tobacco” are not mutually exclusive; therefore, some columns do not add up to the total number of publications reviewed.
<sup>a</sup> GTSS – Global Tobacco Surveillance System
<sup>b</sup> GATS – Global Adult Tobacco Survey
<sup>c</sup> GYTS – Global Youth Tobacco Survey
<sup>d</sup> Six articles used both GATS and GYTS are why the sum of the percentages of GATS and GYTS is more than 100%. <sup>e</sup> Abstract only – records found through our search that were only abstracts for conferences or special issues. <sup>f</sup> Publications were searched only in English, Chinese, French and Spanish. There were zero publications in French found thus we excluded it from the table. <sup>g</sup> Paper published with GTSS data from WHO regions: AFRO – WHO Regional office for Africa, EURO – WHO Regional office for Europe, EMRO - WHO Regional Office for the Eastern Mediterranean, SEARO – Regional office for the South-East Asia Region, PAHO – Pan American Health Organization and WPRO – WHO regional Office for the Western Pacific Region <sup>h</sup> World Bank country income classification <sup>i</sup> Smoked tobacco use included: Smoked tobacco products include cigarettes, cigars, bidis, and kreteks. <sup>j</sup> Smokeless tobacco products included: chewed, sniffed through the nose or held in mouth. <sup>k</sup> Other statistical software used for analysis included: Strata, MLwin, Durbin-Watson, Mplus, AS.

Most publications reported data on smoked tobacco use 94.5% (n=466), while 30.6% (151) reported data on smokeless tobacco use, and 4.1% (n=20) reported data on e-cigarettes. Most publications (97.2%) presented data on both sexes (male and female study participants together), while a small percentage of publications focused their analysis only on females (2.0%, n=10) or males (0.8%, n=4). Publications used several statistical programs to analyze the data, the most used program was SPSS (21.9%, n=108)(Table 1)

Publications by WHO region were 19.7% (n=97) in the AFRO region, 21.5% (n=106) in the EMRO, 24.3% (n=120) in PAHO, 27.2% (n=134) in the EURO region, 28.8% (n=142) in the WPRO region, and 42.6% (n=210) in SEARO region. The highest number of publications for GATS surveillance data was in the SEARO region (n=136) and the lowest number in the AFRO region (n=29). For GYTS data, the number of publications was more evenly distributed across regions with the lowest percentage of publications being surveillance data from PAHO (n=52) and the highest being data from SEARO (n=77). (Table 1)

All countries that had conducted at least one round of GATS or GYTS had at least one publication (Fig 2). Twelve countries in the map (Bangladesh, China, Egypt, India, Mexico, Philippines, Poland, Russian Federation, Thailand, Turkey, Ukraine, and Vietnam) had GTSS data published more than 50 times, while 56 countries had 1 to 5 publications (refer to Supplemental Table 1 to see a full list of publications using GTSS by country).

We found that overall, the percentage of publications that focused on *Monitor* (M) was the highest with 37.3% of publications, and the MPOWER topic with the least number of publications was *Raise* (R) 7.2%. In all WHO regions, M was the topic that was published about most while R was the topic published about least. This pattern was similar in all WHO regions (Fig 3).

### Lead author contribution to the GTSS literature

Of the total number of publications that met the criteria, 37.1% (n=183) of publications were led by authors from the PAHO region, followed by authors from SEARO 27.1%(n=107), EURO 15.4% (n=76), WPRO 13.6% (n=67) and AFRO 7.1% (n=35) and EMRO 5.1% (n=25). The

PAHO and SEARO regions had the highest production of publications where the lead author’s origin was the same as the country where the publication originated from. However, we performed an outlier analysis which showed PAHO without the United States and WHO South-East Asia without India. This analysis showed PAHO without the United States at 9.7% (*n* = 48) lead author publications and WHO South-East Asia without India at 4.5% (*n* = 22).. (Table 1) After classifying lead author country by World Bank Income Classification (31), most GTSS publications (46.4%, n=228) were led by authors in h-ICs, while u-MICs produced 20% of the research (n=99), l-MCs produced 31.6% (n=155), and l-ICs accounted for 1.8% (n=9) of the research productivity (Table 1).

When looking at authorship by country, over 50% of the lead author research output was led by five countries: the United States (27.4%, n=135), followed by India (16.8%, n=83), Malaysia (4.5%, n=22), Vietnam (4.1%, n=20) and Poland (3.2%, n=16) (Fig 4). However, when juxtaposing our findings of research capacity with tobacco use burden, capacity does not match the tobacco burden. Jordan, Greece and Bangladesh have one of the highest adult tobacco use reported, but have a total of four (0.8%), five (1.0%) and ten (2.0%) GTSS lead author publications, respectively. Lead authors in India and the U.S. had the highest number of publications compared to the rest of their whole WHO regions.

## Discussion

This study is the first to look at GTSS publication output, origin of GTSS data as compared to origin of the lead author for the paper and patterns of publication by country, world region and income.

Countries with high burden of non-communicable diseases and risk factors (such as physical inactivity) tend to have lower research productivity.(2, 32) Our review identified unequal distribution of lead authorship and tobacco burden.. Over 80% of the world’s 1.3 billion tobacco users live in low- and middle-income countries(12) and we found a lack of peer-reviewed publications using GTSS data that had lead authors from LMIC. This analysis could be driven by the high numbers of tobacco users in just China and India alone. These two countries alone account for more than half of the annual deaths that tobacco causes worldwide each year as they both are Middle Income Countries. Multi-country publications were also included in this analysis, some of which contain tobacco surveillance data from over 130 countries; although all countries who have conducted a GTSS survey were found in our review, most have not published their individual data (only multi-country peer-reviewed publications) that can help advance country-level tobacco control efforts.

Disparities in publication of GTSS data in peer-reviewed publications were noted between regions and countries. PAHO and WHO South-East Asia had the highest percentage of lead authorship, but this was predominantly driven by two countries: the United States in PAHO and India in WHO South-East Asia. The output of publications from lead authors in the U.S. and India combined surpassed the publication of lead authors by the rest of AFRO, EMRO, EURO, and WPRO regions put together. Following India and the U.S., the countries with highest lead authorship were Malaysia, Vietnam, Poland, Mexico, Bangladesh, The United Kingdom, and Zambia. These disparities in publication are not explained by availability of GTSS data. Although the U.S. has not implemented a GTSS survey, the U.S. had the highest number of lead author publications of GTSS data, this could be explained possibly by the funding being based in the U.S.

Another issue that could have explained these differences could be access to GTSS surveillance data, including issues such as internet connectivity to download large files. Theoretically, all researchers with access to the internet have access to all GTSS data through platforms such as https://www.cdc.gov/tobacco/global/gtss/gtssdata/index.html, www.gtssacademy.org, and https://extranet.who.int/ncdsmicrodata/index.php/home. However, internet connectivity might vary by country and explain some of the differences in access to GTSS data. Additionally, researchers have not evaluated global access, availability and awareness of the above websites and resources.

Countries that have a higher prevalence of tobacco use (above 30%) and have implemented GTSS, have low lead authorship. This is in line with previous studies showing the huge burden of some diseases and risk factors in LMIC (such as CVD and physical inactivity) and the disparity in publications (2, 32, 33). Overall, the AFRO region had the lowest number of publications (n=97) but had a relative high number of youth (GYTS) publications (n=69), posing them as a model for other regions to learn about what AFRO is doing for research dissemination efforts among youth. This is very important given that the AFRO region is the fastest growing among the six WHO regions and has been estimated to have a rise in tobacco consumption to 37% by 2025 (15).

Disparities in global health research have been previously described as the 10/90 divide, where less than 10% of the world’s research resources are allocated to 90% of all preventable deaths worldwide (34). A recent analysis of authorship trends in The Lancet Global Health (10) found an under-representation of papers that came from authors in LMICs stating that 35% of the papers in their journal came from authors in LMIC, while 92% of the articles addressed issues in these same countries, which points to the underrepresentation of most LMICs in research literature. Another study from 2004 found similar disparities in contributing authors from LMICs in high five impact journals (11). In our analysis, l-ICs, as defined by the World Bank Income Classification, had 1.8% (9 publications overall) lead authorship, following u-MICs (20.2%, n=99), l-MICs (31.6%, n=155) and h-ICs (46.4%, n=228). L-MICs had a higher lead authorship than u-MICs, and this could be explained by India’s high number of publications, because the country falls under the l-MIC World Bank Classification.

The underrepresentation of tobacco literature from lead authors in LMIC is not a new finding (7, 17, 35), however, the reasons for these disparities could be due to several factors. High skill migration, or “brain drain”, is a documented pattern globally (36); future work could assess if authors from the countries where the data originates have relocated their careers to the U.S., India, or other highly publishing countries. Additionally, resources and workforce capacity to publish research is scarce in LMIC. There may be fewer graduate level training programs available in some regions (i.e., Latin America) (37), resulting in limited numbers of trained local researchers and thereby fewer tobacco-related publications. Journals may be more accessible to authors within highly publishing countries such as the U.S. or India due to the location of institutions hosting journals within these countries. Language limitations should also be considered: many journals require submission of manuscripts in English; capable researchers who do not have familiarity with English may be unable to prepare or submit manuscripts in English, or submissions of native English speakers may be more likely to be accepted for publication in indexed journals. There are disparities in availability of mentorship toward peer-reviewed publication, disparities in capacity to analyze, summarize, and disseminate surveillance data, and disparities in resources available to hire either contracted or institutional analysts (38-40). In the context of competing public health priorities for Ministries of Health in low-resource, low-staff settings, researchers may not have the time or support to write papers for publication. Other competing organizational priorities, such as grant writing or donor reporting, may also decrease time available for manuscript preparation or submission, especially in LMICs where health workforce capacity is already identified as a challenge because of limited number of staff and other competing priorities (41). Lesser academic incentive for publication outside the U.S. and India, if present, may also contribute to publication disparities. If editorial boards of peer-reviewed journals are not of diverse national origin, the composition of persons selecting manuscripts for publication may also contribute to publication disparities (42-44). Fees for publication may contribute to disparities in publication (45), although this does not explain disparately low numbers of publications out of the United Kingdom or other HICs: although some journals waive their fee for publication for LMICs (46, 47), authors may not know this, and may therefore perceive a financial barrier to publication.

In addition to identifying disparities in numbers of lead author publications by country and region, we additionally observed unequal distribution of publication focus by MPOWER measure. When analyzing publications by MPOWER measure of focus, we found that most of the publications that used GTSS data focused on M (Monitor tobacco use and prevention policies) with some focusing also on P (protect people from secondhand smoke) W (warn about the dangers of tobacco smoke) E (Enforce bans on tobacco advertising, promotion and sponsorship) and O (Offer help to quit tobacco use). In our analysis we found few publications that focused on R (Raise taxes on tobacco). The 2021 WHO Global Tobacco Control Report states that “…while being the most effective way to reduce tobacco use, taxation is still the MPOWER policy with the lowest population coverage and has not increased from the 13% achieved in 2018.” (48) This highlights a potential opportunity to expand the focus of scientific publications of GTSS data.

In addition to identification of disparities in publications and unequal utilization of MPOWER measure of publication focus, we additionally observed a paucity of publications addressing new and emerging tobacco products. The 2021 Global Tobacco Control Report highlights the importance of this emerging public health threat and the lack of data and research worldwide for this changing tobacco product landscape (48). In our analysis, we found that only 4.1% (20 publications) of all GTSS publications analyzed data on e-cigarettes, 3.6% (9 publications) for GATS and 4.8 (12 publications) for GYTS. This could be due in part to the e-cigarette questions on GTSS are optional, thus leaving it up to each country to ask about novel and emerging products and that e-cigarettes and emerging new products are relatively new. This may present a challenge to health equity if countries that are likely to see surges in new and emerging products are not equipped to collect the data or to publish their findings.

This study has some limitations. First, our search was limited to manuscripts in English, Spanish, French and Chinese and may be underreporting for publications in other languages. Second, our search strategy only addressed peer-reviewed databases in public health literature, so we did not include publications and reports in the grey literature or other types of peer-reviewed reports like economic publications. Third, the origin of lead authorship reflects the institution where the author was working at the time of publishing and not necessarily where the author is from and hence, we could not capture the actual country of origin of the lead author, (lead authors could be originally from LMICs but may be working or studying in a HICs institution). Fourth, we were only able to collect lead author country data leaving co-authors’ country of origin out, and fifth, we did not focus on an explicit assessment of publication quality.

The Global Tobacco Surveillance System started more than 20 years ago and has been implemented in over 180 countries and locations (49). Since then, there have been several initiatives to strengthen capacity to use and disseminate GTSS and tobacco control data globally including the Data-to-Action Workshops (50) which aim to build capacity to use data to inform and disseminate tobacco prevention and control strategies, the Tobacco Control Scholars Program focusing on peer-to-peer mentoring to publish scientific manuscripts in tobacco control, and established academic training programs like the Field Epidemiology Training Program (51). Having local and current peer-reviewed research could help researchers, decision makers and local organizations prioritize strategies especially as the burden of disease falls in these countries. Some initiatives that have helped enhance capacity for tobacco control have done so by building researchers’ and practitioners’ ability to understand, use, and disseminate tobacco control data among researchers and practitioners. The findings in this bibliometric inventory can be used to identify surveillance needs, research gaps and opportunities for data dissemination as well as prioritize building research capacity and mentoring initiatives to enhance the use of GTSS data in LMICs.

## Data Availability

All related data are included in the manuscript or in the supplementary file.

## Acknowledgements

The authors would like to express our grateful thanks to Christine Shi from the CDC Foundation as she assisted with the translation and extraction of the three papers in Chinese. Partial financial support for this paper was provided by the CDC Foundation with a grant from Bloomberg Philanthropies.

---

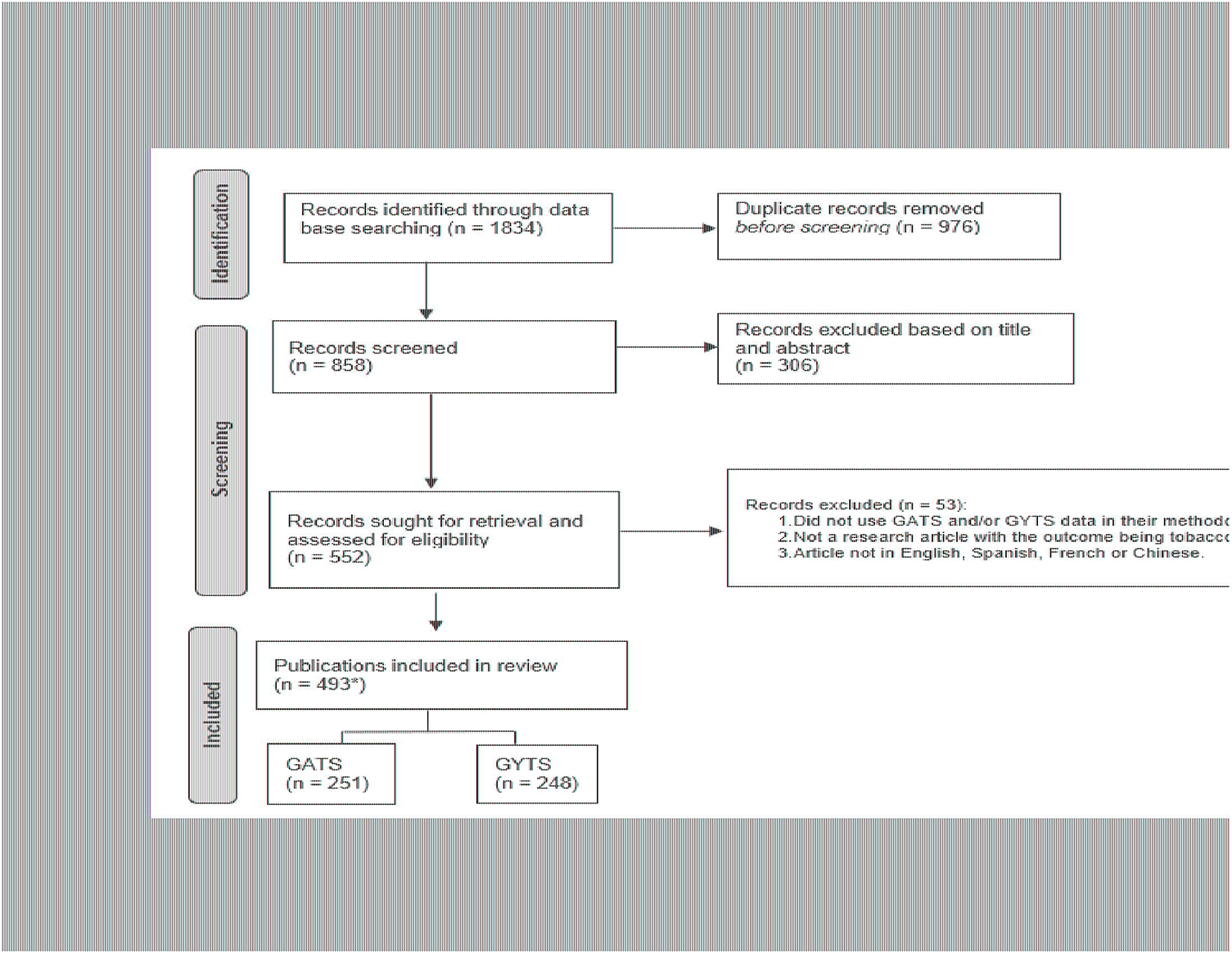

